# Neurostructural associations with traumatic experiences during child- and adulthood

**DOI:** 10.1101/2022.05.20.22275355

**Authors:** Sebastian Siehl, Maurizio Sicorello, Julia Herzog, Frauke Nees, Nikolaus Kleindienst, Martin Bohus, Meike Müller-Engelmann, Regina Steil, Kathlen Priebe, Christian Schmahl, Herta Flor

**Author notes:** **Corresponding author**: Sebastian Siehl, Ph.D., Institute of Medical Psychology and Medical Sociology, University Medical Center Schleswig-Holstein, Kiel University, Preußerstrasse 1-9, 24105 Kiel. shared first authorship (Sebastian Siehl, Maurizio Sicorello). **Previous Presentation:** Siehl, S. (2021). Structural and functional neuroplasticity in posttraumatic stress disorder: fear learning and context processing. [doctoral thesis, University of Mannheim]. https://madoc.bib.uni-mannheim.de. **Data availability statement:** We report how we determined our sample size, all data exclusions, all inclusion/exclusion criteria, whether inclusion/exclusion criteria were established prior to data analysis, all manipulations, and all measures in the study. Ethical restrictions to protect participant confidentiality prevent us from making study data publicly available. This also refers to the analysis/experimental code, and any other digital materials, where participant-related information (like sex or psychopathological status) are also included. Readers seeking access to the study data and materials should contact the corresponding author based on a formal collaboration agreement. This formal collaboration agreement indicates that data will be shared with other researchers who agree to work with the authors, and for the sole purpose of verifying the claims in the paper. The data and materials will be released to requestors after approval of this formal collaboration agreement by the local Ethics Committee of the Medical Faculty Mannheim.

## Abstract

**Background:** Adverse experiences can lead to severe mental health problems such as posttraumatic stress disorder (PTSD) throughout the lifespan. In individuals with PTSD, both global and local brain volume reductions have been reported—especially in the amygdala and hippocampus—while the literature on childhood maltreatment suggests strong dependency on the timing of adverse events. In the present study, we pooled data from two studies to contrast effects of reported trauma-exposure during neurodevelopmentally sensitive periods in early life with trauma-exposure during adulthood.

**Methods:** A total of 155 women were allocated into one of six age-matched groups according to timing of traumatization (childhood vs adulthood) and psychopathology (PTSD vs trauma-exposed healthy vs trauma-naïve healthy). Volumes of amygdala and hippocampus were compared between these groups. Six additional exploratory regions of interest (ROI) were included based on a recent meta-analysis.

**Results:** Amygdala volume was strongly dependent on timing of traumatization: Smaller amygdala volumes were observed in the childhood sample, while larger volumes were observed in the adulthood sample. Hippocampal volume comparisons revealed no statistically significant differences, although the descriptive pattern was similar to that found for the amygdala. The remaining exploratory ROIs showed significant group effects, but no timing effects.

**Conclusions:** Timing of traumatization was associated with amygdala volumes throughout the lifespan, with opposite effects dependent on age at trauma occurrence. The relevance of potential confounders like trauma-type and multiplicity is discussed.

## Introduction

Posttraumatic stress disorder (PTSD) is a debilitating condition affecting about 3.9% of the global population during their lifetime (1). It is characterized by intrusive re-experiencing of traumatic events, avoidance of trauma-related memories and external cues, alterations in cognition, mood, arousal, and reactivity (DSM-5; American Psychiatric Association, 2013). Motivated by its severe consequences for well-being, health, and mortality (3,4) and the extremely high prevalence of traumatic experiences worldwide (70% lifetime prevalence; Kessler et al., 2017), there have been major ongoing efforts to identify vulnerability factors and refine pathophysiological models of PTSD with a strong focus on neuroimaging.

Among the most consistent neuroimaging findings are lower regional and global white- and grey-matter brain volume in PTSD patients (6–8). In terms of local regions, most research has been devoted to the amygdala and the hippocampus. Both regions are involved in cued and contextualized fear learning, show relative consistent volume reductions in PTSD samples, and exhibit stress-dependent alterations in animal studies (9–12). Moreover, smaller local volumes have been reported for the insula and the medial prefrontal cortex (PFC) (6), including alterations in interhemispheric white matter tracts in the PFC (13). These regions play a key role in psychobiological models of PTSD (11,14).

For correct interpretation of these findings, it is crucial to distinguish which neural alterations are functionally related to PTSD symptoms and not a mere consequence of stress-exposure in the absence of mental or physical sequelae (15). A meta-analysis by Paquola and colleagues (16) demonstrated that hippocampal atrophies can be found even in healthy stress-exposed samples, while amygdala atrophies were only present in samples with PTSD. Using a more complete approach, the meta-analysis by Bromis and colleagues (6) found that PTSD samples had smaller hippocampal volumes than trauma-exposed controls, which in turn had smaller volumes than trauma-naive controls, potentially reflecting a dose-response relationship of stress exposure. A similar pattern was descriptively found for the amygdala, but differences between groups were smaller and only statistically significant when the PTSD group was compared to the pooled control groups.

A major challenge for the field is the potential dependency of stress-brain associations on the timing of adverse experiences. This challenge has received substantial research attention during recent years in the literature on adverse childhood experiences, which is one of the strongest risk factors for PTSD (5). Volume reductions following childhood maltreatment for both hippocampus and amygdala appear to be dependent on the developmental timing of events, supporting the existence of sensitive neurodevelopmental periods (17–22). Moreover, first evidence indicates timing-effects generalize to amygdala function, revealing differential effects of trauma exposure and PTSD (23,24). These studies on trauma timing have added nuance to the interpretation of neural markers and contribute to theories of (mal-) adaptive neurodevelopment. Nevertheless, they have thus far focused on the period of childhood and adolescence, while studies on stress-exposed adults suggest volumetric alterations can still emerge later in life, although it is unclear whether these include the amygdala and hippocampus (25).

In the present study, we aimed to contrast the neurostructural associations with early and late trauma-exposure, while also accounting for the role of psychopathology. We compared regional brain volumes of women who (a) either experienced traumatic events before or after entering adulthood (i.e., age 18) and (b) either developed PTSD or remained physically and mentally healthy. A trauma-naive healthy control group was included as well to assess the general effect of trauma exposure. All groups were matched for age to avoid confounding (26). The main focus of our study was on the amygdala and the hippocampus, which have by far the strongest theoretical and empirical basis for associations between trauma timing and psychopathology. For exploratory analyses, we further included all structures for which differences between PTSD and (combined) controls were reported in a previous meta-analysis (6) to provide a first basis for the investigation of trauma timing effects on these regions. These exploratory regions included the inferior fronto-orbital gyrus (IFOG), anterior cingulate gyrus (ACG), anterior insula, posterior insula, middle temporal gyrus (MTG), and superior frontal gyrus (SFG).

## Methods and Materials

### 2.1 Participants

The total sample of 156 adult women (mean age = 35.3; SD = 10.6; range 20-60 years) was pooled from two cross-sectional MRI studies on adverse experiences and psychopathology, conducted at the same scanner and facilities between 2010 and 2018 at the Central Institute of Mental Health (CIMH) in Mannheim, Germany. One participant had to be excluded from the analyses due to motion artifacts, resulting in an effective sample size of 155 participants. Sample 1 assessed adult women with traumatic experiences before the age of 18; sample 2 assessed adult women with traumatic experiences during adulthood. Both studies comprised three groups: patients with trauma-exposure and PTSD (PTSD), trauma-exposed healthy controls (TC), and trauma-naive healthy controls (HC). Hence, the pooled sample consists of six groups with 26 female participants in each group (see 2.2, 2.3 for detailed description). Groups from the childhood sample are denoted with a subscripted “child” (e.g., PTSD_child_); groups from the adulthood sample are denoted with a subscripted “adult” (e.g., PTSD_adult_). Demographic and clinical data can be found in *Table 1* and *Suppl. Table 1*. For further notable differences between the two samples, see the methods section on procedures and the discussion section on limitations.

**Table 1.**
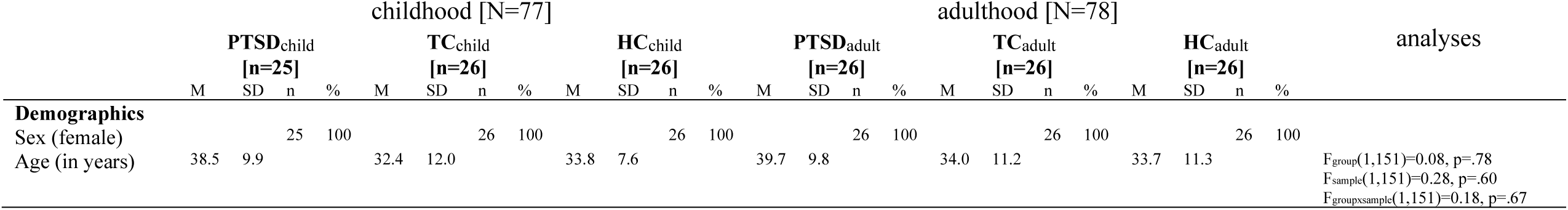
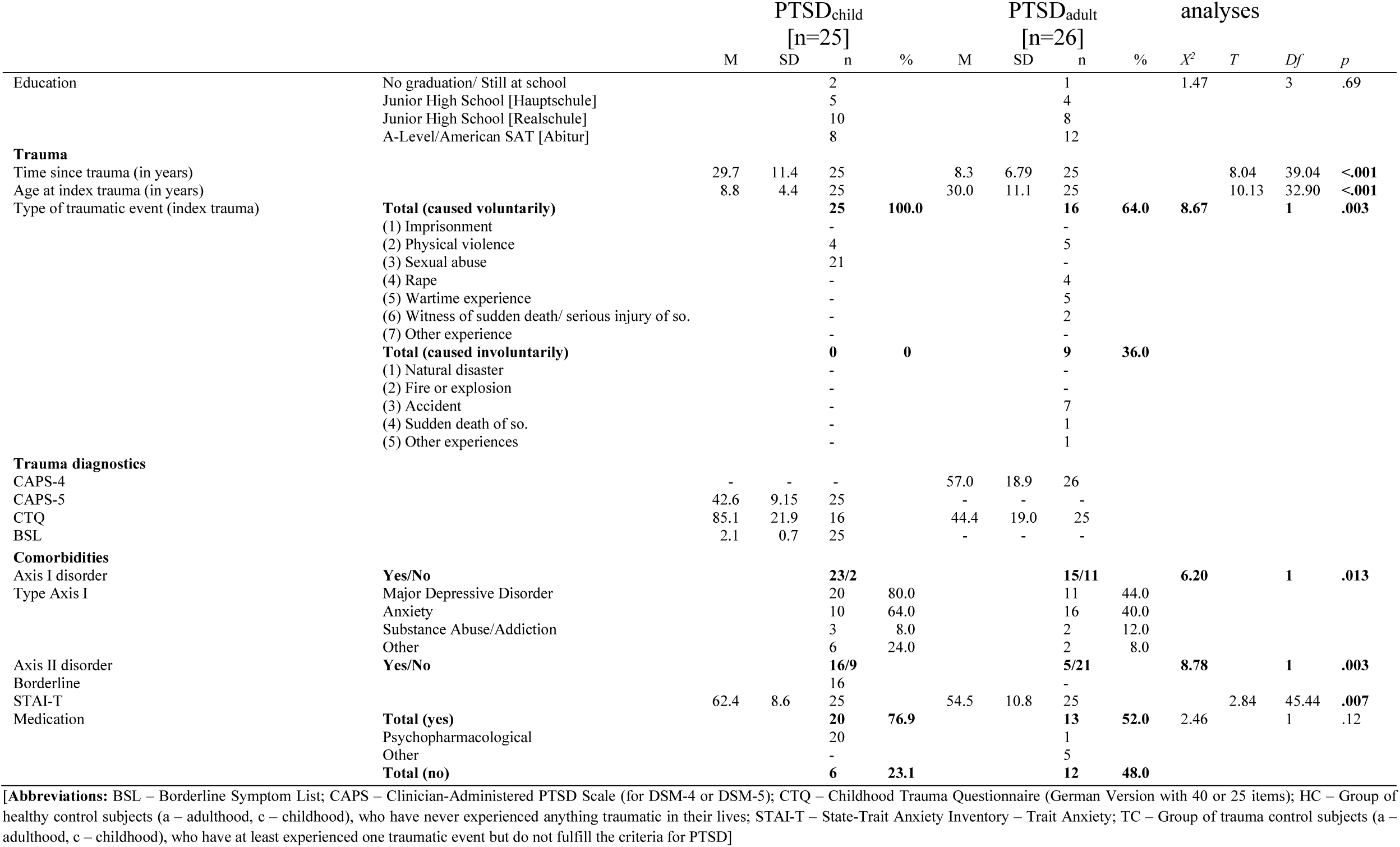
Socio-demographic and clinical characteristics of sample.

All participants received reimbursement for participation (10€/h) and travel expenses. Patients were offered treatment in the outpatient clinics of the CIMH in Mannheim and the outpatient treatment center of Goethe University in Frankfurt. The study was carried out following the Code of Ethics of the World Medical Association (World Medical Association, Declaration of Helsinki, seventh revision, 2013). The study was approved by the Ethical Review Board of the Medical Faculty Mannheim (Heidelberg University) and the ethics committee of the Goethe University. All participants gave written informed consent including consent for data re-analysis.

### 2.2 Procedures

#### 2.2.1 Sample 1: Trauma experience in childhood

Participants with PTSD after traumatic experiences in childhood (PTSD_child_) were recruited from a larger randomized controlled psychotherapy study (27,28). Inclusion criteria were the experience of physical or sexual abuse before the age of 18 as well as female sex and gender identity. Moreover, participants had to fulfill at least 3 criteria for Borderline Personality Disorder (BPD), including the criterion for affective instability. They underwent MRI measurements between randomization and the first therapy session. PTSD was assessed by trained diagnosticians using the Structured Clinical Interview for DSM-IV Axis I Disorders (SCID-I; (29)). Trauma exposure was measured by the Life Events Checklist (LEC-5; (30)), which was also used to determine the index trauma. Additionally, the Clinician Administered PTSD Scale for DSM-5 (CAPS-5; (31)) and the BPD section of the International Personality Disorder Examination were administered (IPDE; (32)). The CAPS-5 assesses the severity of 20 symptoms in relation to the index trauma. Symptoms are assessed on a 5-point scale ranging from 0 (no impairment) to 4 (extreme impairment). In addition to establishing PTBS diagnoses, the total CAPS-5 score, with a maximum of 80, gives an indication of clinical severity.

Further self-report measures included retrospective questionnaires on childhood trauma (Childhood Trauma Questionnaire [CTQ]; (33)), PTSD symptoms (PTSD checklist for DSM-5 [PCL-5]; (34); Davidson Trauma Scale [DTS]; (35)), and severity of depressive mood (Beck Depression Inventory 2 [BDI-II]; (36)). Healthy trauma-exposed controls (TC_child_) who reported physical or sexual abuse before the age of 18 and healthy trauma-naive controls (HC_child_) were recruited with advertisements in local newspapers, flyers and over the internet.

Exclusion criteria for all participants were age under 18 or over 65, metal implants, pregnancy, left-handedness, and claustrophobia. Exclusion criteria for PTSD participants specifically covered current and lifetime schizophrenia or bipolar-I disorder, mental retardation, or severe psychopathology requiring immediate treatment in a different setting (e.g., BMI<16.5), medical conditions contradicting exposure-based treatment (e.g., pregnancy), a highly unstable life situation (e.g., homelessness), a life-threatening suicide attempt within the last two months, and substance dependence with no abstinence within two months prior to the study. Exclusion criteria for the trauma controls were any current or previous mental disorder, any prior psychotherapy, or any intake of psychotropic medication.

Structural MRI analyses on a partially overlapping sample have been previously published (18).

#### 2.2.2 Sample 2: Trauma experience in adulthood

All participants were assessed by a trained psychologist for trauma exposure using a list of possible traumatic events, taken from the Posttraumatic Diagnostic Scale (37), followed by the SCID-I and II for DSM-IV-TR (29,38,39). Participants were assigned to the PTSD group, when the diagnostic criteria were fulfilled in the SCID-I interview. The index events reported by participants in sample 2 were not exclusively limited to interpersonal violence. Participants, reporting other traumatic events fulfilling DSM-V criteria A of the PTSD diagnostics, were also included. In addition, participants were assessed with the German version of the Clinician-Administered Posttraumatic Stress Scale for DSM-IV (CAPS; (40,41)) and had to fulfill criteria B through F. The CAPS score for symptom severity ranges from 0 to 100, assessed on a 5-point scale ranging from zero (“never”/ “none) to four “most or all the time”/ “extreme”).

For the sample of patients with trauma experience in adulthood (PTSD_adult_) the following exclusion criteria were applied in the original studies: younger than 18 years, any traumatic experience (interpersonal or any other) before the age of 18 years, comorbid current or lifetime psychotic symptoms, current alcohol/ drug dependence or abuse, borderline personality disorder, cardiovascular or neurological disorders, brain injury, acute pain, continuous pain or medication for attention deficit hyperactivity disorder, pregnancy and metal implants. Importantly, patients and trauma-exposed individuals in sample two had no traumatic experience before the age of 18 years (telephone screening with PDS and SCID). The healthy trauma-exposed individuals in this sample were trauma-exposed in adulthood (TC_adult_) but did not fulfil any criteria for a current or past mental disorder as assessed with the SCID-Interview as well as the CAPS. Healthy trauma-naive individuals (HC_adult_) did not fulfil any criteria for a mental disorder.

### 2.4 MRI Data acquisition

For both samples, we acquired T1-weighted, magnetization-prepared, rapid-acquisition gradient echo (MPRAGE) images using the same 3T Magnetom TRIO whole body magnetic resonance scanner (Siemens Medical Solutions, Erlangen, Germany) equipped with a standard 12-channel volume head coil. Slightly different acquisition parameters were used in each sample, for which we accounted in the preprocessing steps. In the sample of trauma experience in childhood (sample 1), the following parameters were applied: TR = 1570 ms, TE = 2.75 ms, flip angle 15°, FOV: 256 ×256 mm^2^, matrix size: 256 × 256, voxel size: 1.0 × 1.0 × 1.0 mm^3^, 176 sagittal slices. In the sample of trauma experience in adulthood (sample 2), the following parameters were applied: TR = 2300 ms, TE = 2.98 ms, flip angle 9°, FOV: 256 ×256 mm^2^, matrix size: 256 × 256, voxel size: 1.0 × 1.0 × 1.1 mm^3^, 160 sagittal slices.

### 2.5 Clinical assessments for both samples

Traumatic childhood experience was assessed with the German version of the Childhood Trauma Questionnaire (CTQ; (42,43)). The self-report instrument assesses the severity of trauma exposure, such as emotional abuse and neglect, physical abuse and neglect as well as sexual abuse. The 25 items ask how often each event occurred during the participant’s upbringing and each item is rated on a 5-point Likert scale ranging from 1 (“never at all”) to 5 (“very often”). The overall sum score was calculated, which is calculated by the sum of the five subscales, ranging from 25 to 125. In sample two, the 40-item version of the CTQ was used, with additional two subscales and six items, in which participants could rate the age in which the childhood experiences occurred. However, for the purpose of this study, we only calculated the sum score of the same 25-items as for sample one.

Trait anxiety was assessed with the German version of the trait-version of the State-Trait-Anxiety-Inventory (STAI-T; (44)), a self-report questionnaire with 20 items, assessed on a 4-point Likert scale ranging from one (“not at all”) to four (“very much”). The total STAI-T scores ranges from 20 to 80, with higher scores being associated with higher levels of trait anxiety.

### 2.6 Pooling and matching of data

The data of the two samples were pooled in a multi-step process. The original pool consisted of over 297 participants including 104 male participants. Male participants were excluded since the sample of traumatic experience in childhood consisted only of female participants. In a next step, data was assessed for completeness (excluding 10 participants). Patients from either of the two samples (childhood and adulthood) were age-matched and in a second step each group of patients was then age-matched to individuals from the TC and HC group (exclusion of 15 participants) manually minimizing the age difference between matched groups. If a patient could be age-matched equally well to a participant from either control group, the participant from the control group was taken with a total intracranial volume (TIV) more similar to the patient’s TIV (exclusion of 12 participants, 1-3 participants from each group).

### 2.7 MRI Preprocessing

The T1-weighted images were preprocessed using the Computational Anatomy Toolbox (CAT12; http://www.neuro.uni-jena.de/cat) on Statistical Parametric Mapping (SPM12; Wellcome Department of Imaging Neuroscience, London, UK) implemented in customized scripts in MATLAB R2016a (The MathWorks Inc., Natick, MA, USA). The preprocessing steps included spatial registration, segmentation into gray and white matter and CSF as well as bias correction of intensity non-uniformities following our previous study (13). We chose the Neuromorphometic atlas (provided by Neuromorphometrics, Inc., MA, USA; http://www.neuromorphometrics.com) for the definition of region of interests (ROIs). We then extracted gray matter volume (in cm^3^) for eight predefined ROIs, following the results by a recent meta-analysis (6): amygdala, hippocampus, IFOG, anterior cingulate ACG, anterior insula, posterior insula, MTG, SFG. Data was assessed for head motion, excluding one participant (from the PTSD_child_ group) moving more than the maximum translation of 1 mm in x-, y-, or z-direction and the maximum angular motion of 1° throughout the course of the scan.

### 2.8 Statistical analyses

Statistical analyses were performed in R-Statistics (45) using the packages dplyr (46) for data processing, and rstatix and emmeans for the analyses and ggplot2 (47) for plotting. We assessed all data for the appropriate assumptions, including normal distribution and outliers. Data was in line with these assumptions, if not stated otherwise below. For the socio-demographic data, two-sample t-tests as well as Chi-square tests of frequency distributions were applied. We then performed a mixed 3 (*group*_*between-subject*_: PTSD, TC, HC) x 2 (*sample*_*between-subject*_: childhood, adulthood) x 2 (*hemisphere*_*within-subject*_: left, right) Analysis of Covariances (ANCOVAs) for each of the eight ROIs with total intracranial volume (TIV) as covariate. To counter inflation of Type I errors, Bonferroni corrections were applied (α/8 = .05/8 = .00625). Post-hoc single-step multiple comparison t-tests were applied with Bonferroni corrections.

## 3 Results

### 3.1 Sample descriptions

The two patient groups (PTSD_child_, PTSD_adult_) had similar education levels (*Table 1, Suppl. Table 1*). The age at index trauma was significantly lower in PTSD_child_ than PTSD_adult_. Similarly, CTQ scores differed strongly between the two groups (*Table 1*). There were significant differences in the types of traumatic events experienced in each patient group, with PTSD_child_ experiencing significantly more interpersonal trauma (e.g., physical or sexual abuse) than PTSD_adult_. Significantly more patients in the PTSD_child_ group had comorbid mental disorders on axis-I as well as borderline personality disorder (Suppl. Table 1). In addition, patients in the PTSD_child_ group reported significantly higher anxiety scores on the STAI-T. Finally, there was no difference in overall number of participants taking medication (dichotomous: yes/no) and kind of medication taken between the PTSD_child_ and PTSD_adult_ groups (Table 1, Suppl. Table 1). Medication dosage was not assessed.

### 3.2 Amygdala

Comprehensive inferential statistics for all regions of interest are reported in Table 2. Region-wise means and standard deviations can be found in Suppl. Table 2.

**Table 2.**
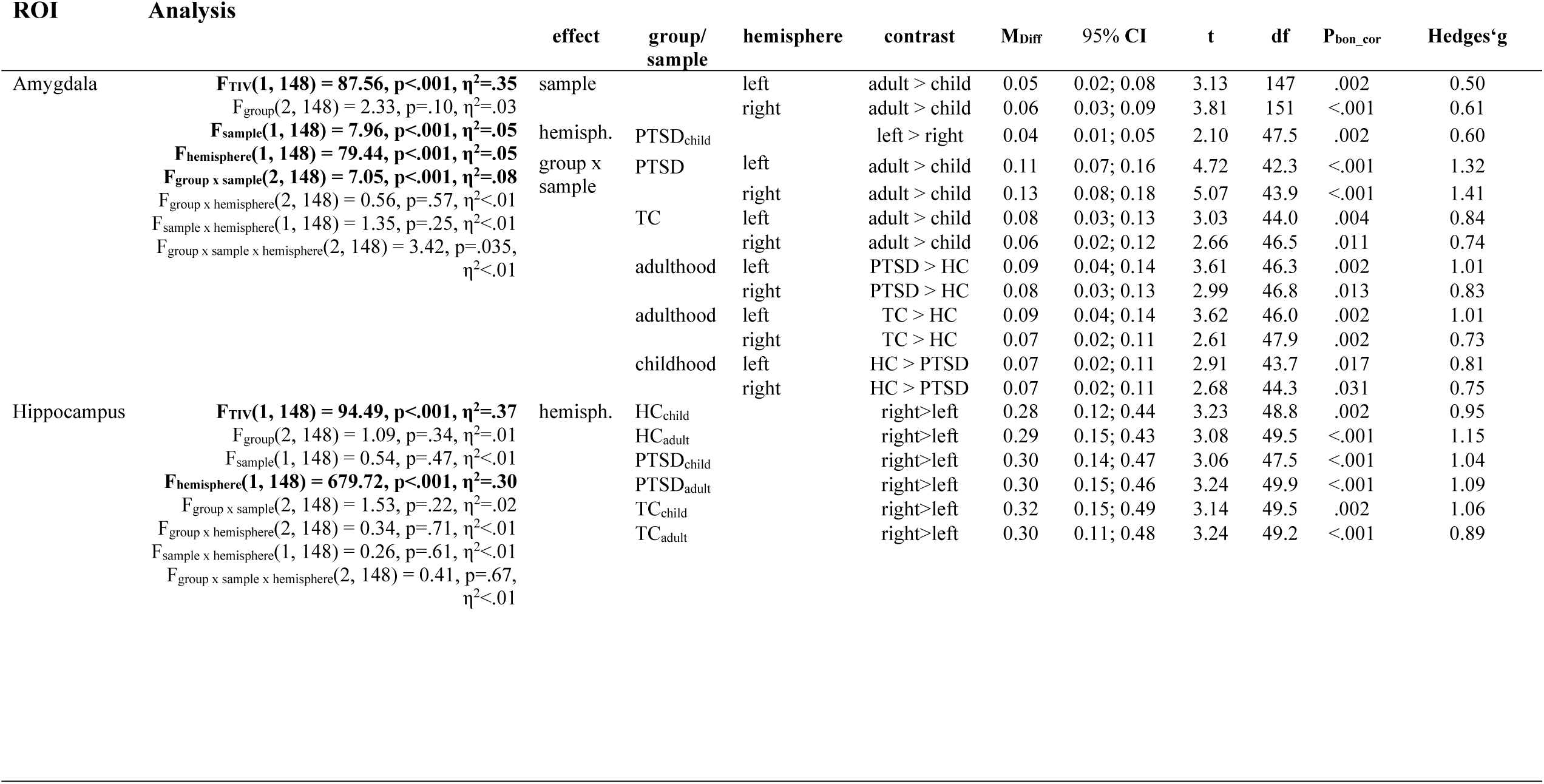
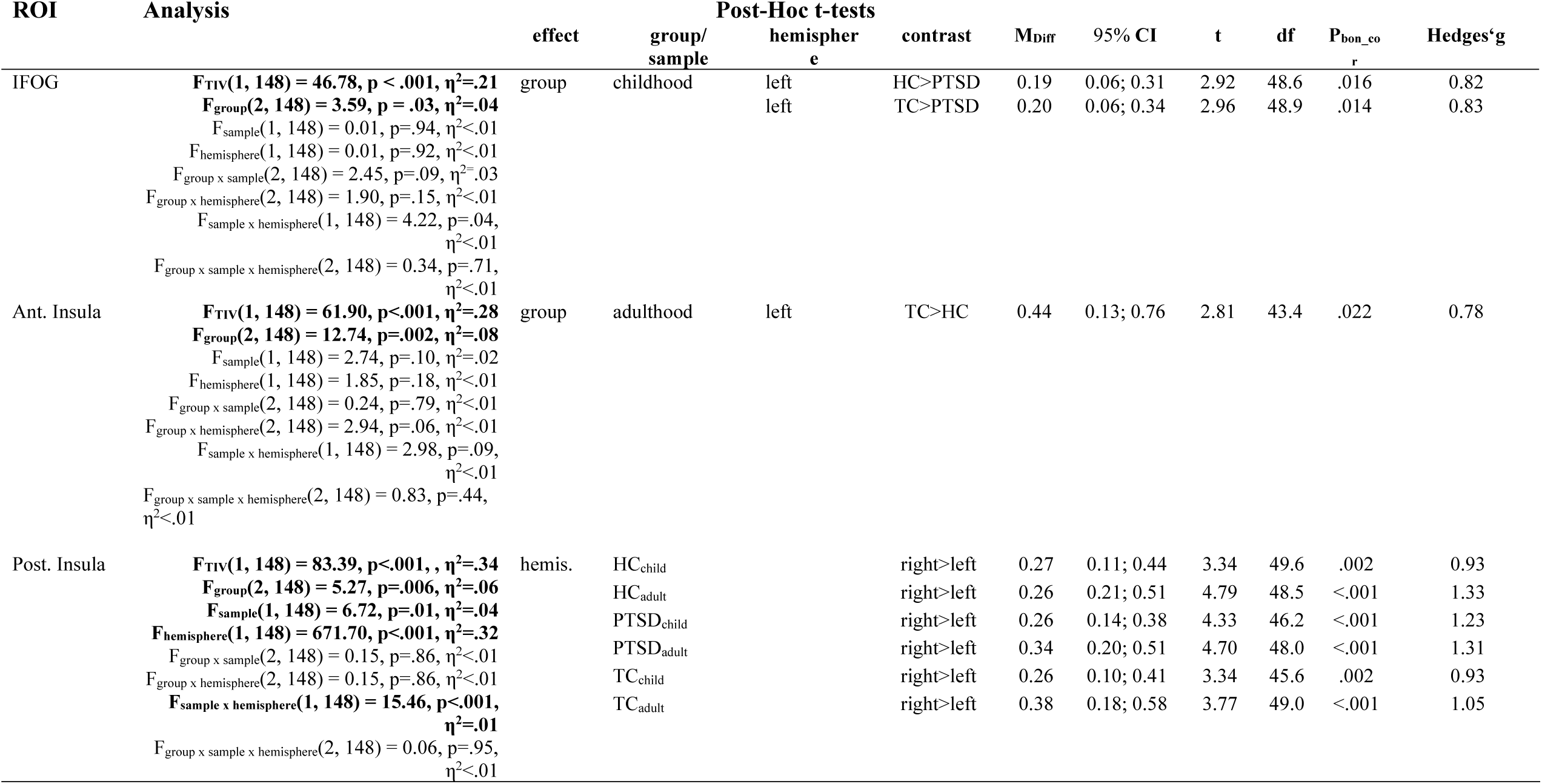
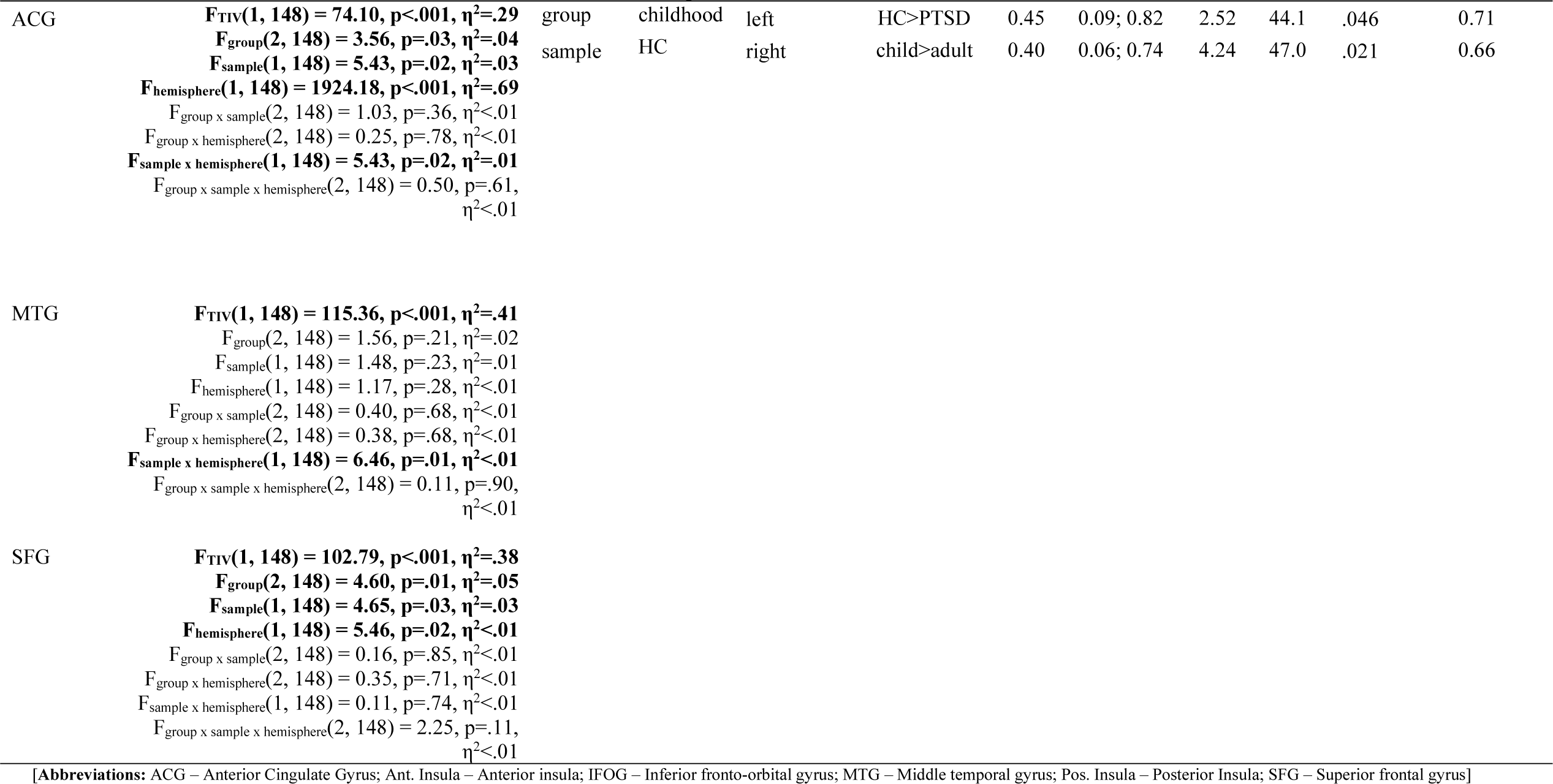
Results of ANCOVAs on volumetric data.

We found a significant interaction of group and sample (*Figure 1; Table 2; Suppl. Table 2*). Post-hoc t-tests revealed an effect of trauma timing: Amygdala volume was significantly higher for participants with adult trauma compared to those with childhood trauma. This was also apparent in a time-series of index traumas in the PTSD groups with a finer time resolution (*Fig.2*, *Suppl. Table 4*). In comparison to the trauma-naive healthy control groups, we found opposite effects dependent on timing: For childhood trauma, the PTSD_child_ group exhibited significantly smaller amygdala volumes than the HC_child_ group. For adulthood trauma, both PTSD_adult_ and TC_adult_ had significantly larger amygdala volumes compared to the HC_adult_ group. We found a significant main effect of hemisphere, with the right amygdala showing significantly lower volume than the left amygdala. There were no further interactions between hemisphere and the other independent variables.

**Figure 1.**
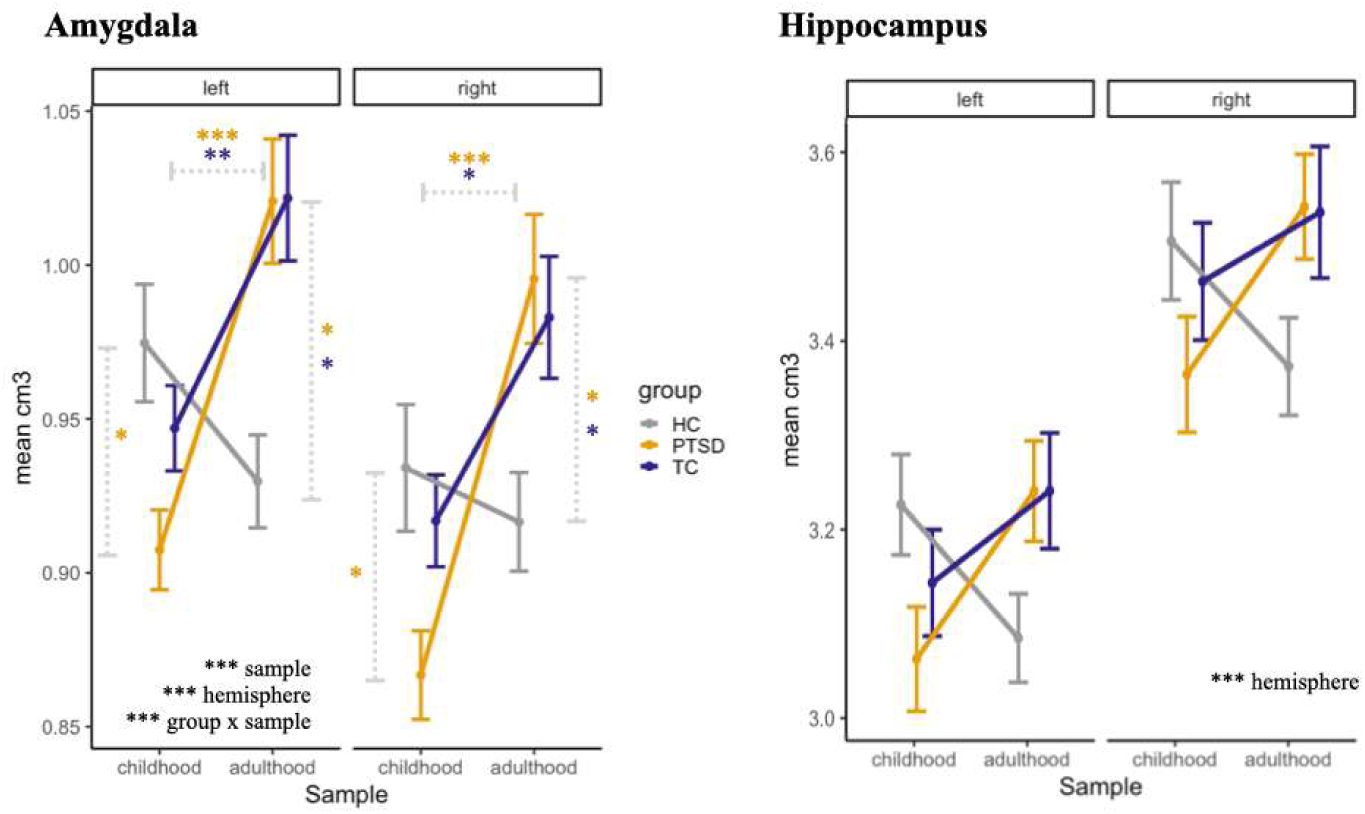
Volumetric differences in the amygdala and hippocampus between samples (childhood, adulthood), groups (PTSD, TC, HC) and hemispheres (left, right) in cm^3^.

**Figure 2.**
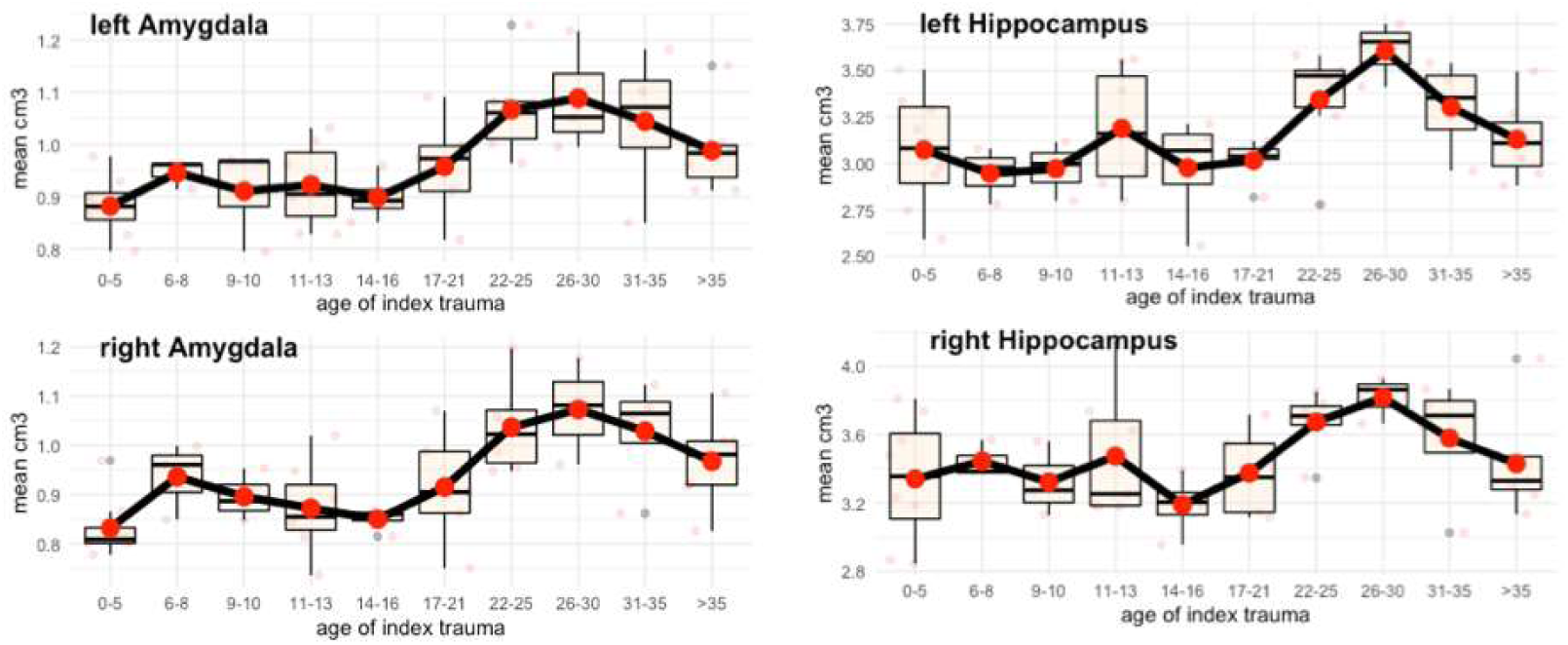
Volumetric differences in the amygdala and hippocampus for both patient groups (PTSDadult, PTSDchild) in time bins defined by the age of the index trauma separately for each hemisphere (left, right) in cm^3^.

### 3.3 Hippocampus

We found a significant main effect of hemisphere, with larger volume in the right hippocampus (*Figure 1; Table 2; Suppl. Table 2*). All remaining effects were not significant. Descriptively, similar to the pattern for the amygdala, only the PTSD_child_ group had smaller hippocampal volumes than their reference groups, while both groups with adulthood trauma actually had slightly *higher* hippocampal volumes, opposite to the expected effect direction.

### 3.4 Exploratory regions of interest

Quantitative results and graphical representations for all exploratory ROIs can be found in *Table 2* and *Figure 3 and 4* (*as well as Suppl. Table 2*).

**Figure 3.**
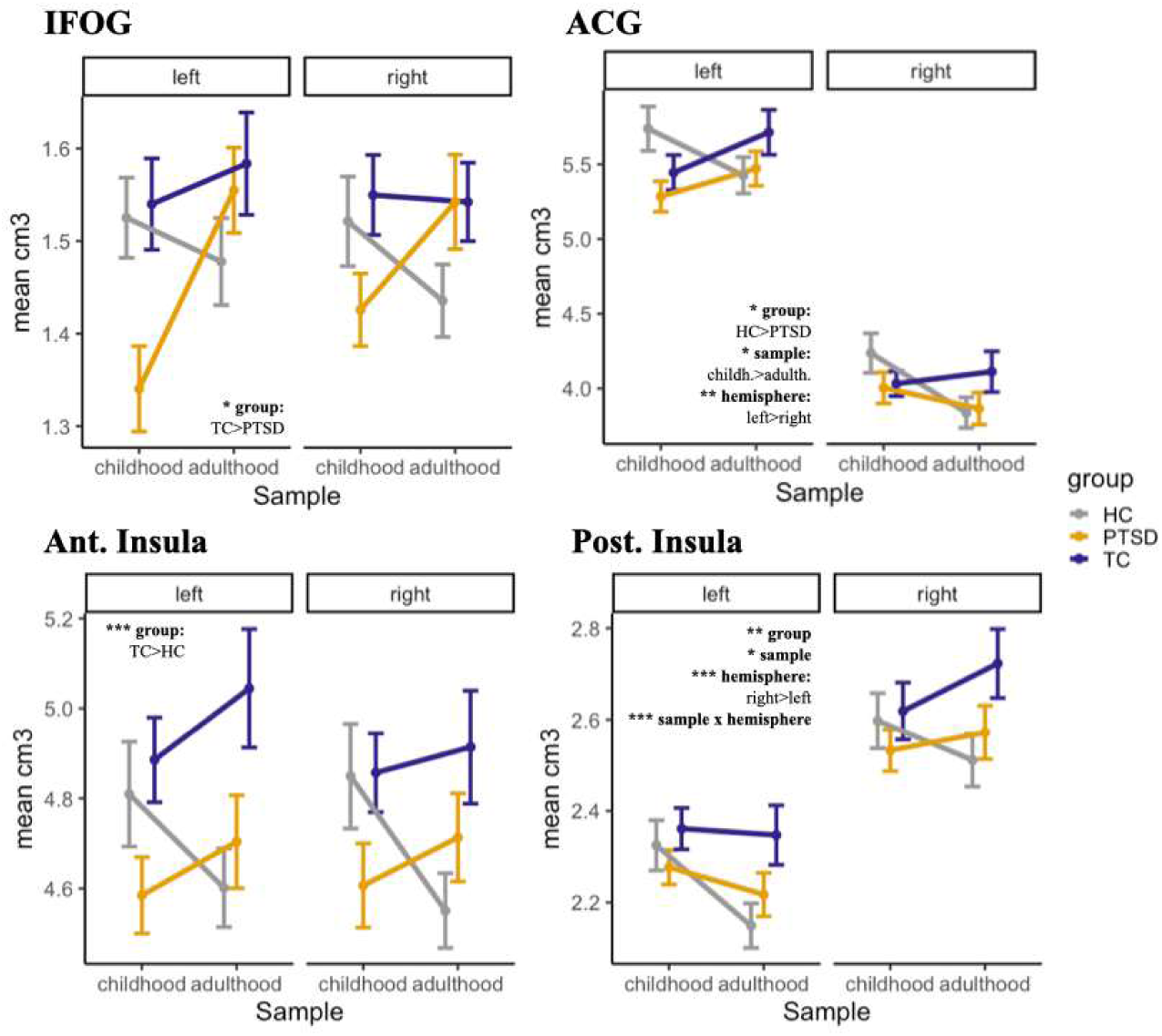
Volumetric differences in the inferior fronto-orbital gyrus (IFOG), anterior cingulate gyrus (ACG), anterior (ant.) and posterior (pos.) insulae between samples (childhood, adulthood), groups (PTSD, TC, HC) and hemispheres (left, right) in cm^3^.

**Figure 4.**
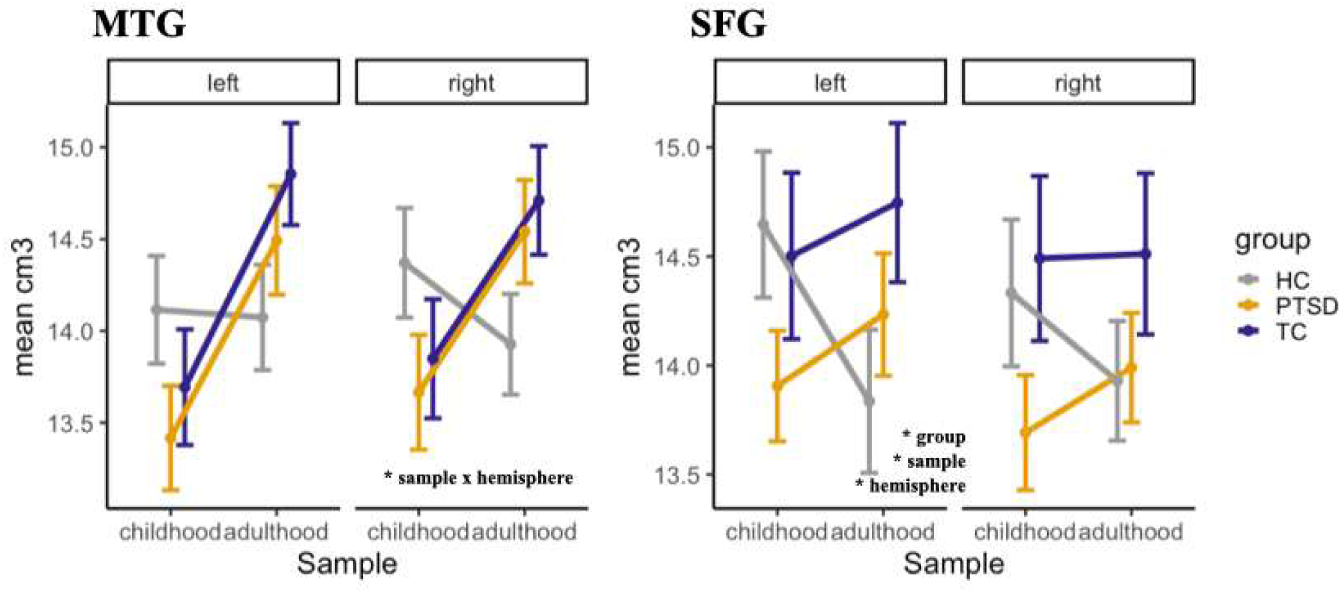
Volumetric differences in the middle temporal gyrus (MTG) and superior frontal gyrus (SFG) between samples (childhood, adulthood), groups (PTSD, TC, HC) and hemispheres (left, right) in cm^3^.

#### 3.4.1 IFOG

A significant main effect of group was found. Post-hoc tests revealed that within the childhood sample, both control groups had larger left IFOG volumes than the PTSD group.

#### 3.4.2 Anterior insula

A significant main effect of group was found. Post-hoc test showed that within the adulthood sample, TCs had larger volumes than HCs.

#### 3.4.3 Posterior insula

There were significant main effects for group, sample, and hemisphere, as well as a significant interaction between sample and hemisphere. Only the post-hoc tests for hemisphere survived multiple comparison correction, confirming larger volumes of the right insula in all groups.

#### 3.4.4 ACG

As for the posterior insula, we found a significant main effect of group, sample, and hemisphere, as well as a significant interaction between sample and hemisphere. Post-hoc effect indicated larger brain volumes in HC_child_ than PTSD_child_ within the left ACG. There was also a significant difference between the two healthy control groups within the right ACG.

#### 3.4.5 MTG

There was a significant interaction between sample and hemisphere, but post hoc tests did not survive correction for multiple comparisons. Descriptively, the significant interaction is most likely driven by the larger right volumes in the two trauma groups exposed during adulthood, with visual similarity to the disordinal pattern found for the amygdala.

#### 3.4.6 SFG

Significant main effects for group, sample, and hemisphere were found, with no post-hoc contrasts surviving correction for multiple comparisons.

## 4 Discussion

In the research literature on early adversity, trauma-induced differences in brain volume are increasingly viewed as largely dependent on the neurodevelopmental timing of events. Still, most studies on sensitive periods limited their scope to events occurring during childhood and adolescence. Extending research on sensitive periods to adverse events during adulthood may further help differentiate early neurodevelopmental processes from life-long plasticity.

The amygdala and the hippocampus play a key role in psychobiological models of PTSD and have been highlighted in research on sensitive periods during early childhood and early adolescence (11,18,48). Building on this research, we found evidence that amygdala volumes strongly depended on the timing of events, revealing qualitative differences between individuals who were traumatized in childhood or adulthood (see the limitation section for a discussion of potential confounders). While participants with PTSD following childhood trauma had *lower* amygdala volumes compared to trauma-naive healthy controls, participants with adult trauma had *higher* amygdala volumes. These *higher* volumes were apparent in both trauma-exposed groups with- and without psychopathology, potentially indicating general neuroplastic events in response to exposure, rather than clinically meaningful differences.

We did not find significant timing effects on hippocampus volumes, which might be due to limited statistical power. Nevertheless, we find it notable that in the groups with trauma-exposure during adulthood, hippocampus volume was descriptively *larger* than in the trauma-naive controls for both hemispheres. This difference did not survive bonferroni-correction, albeit confidence intervals for this comparison were clearly separated. Qualitatively, this descriptive pattern did not show the often reported general reduction in hippocampus volume, but rather matched the pattern found for the amygdala.

In sum, these data agree with the notion that stress-dependent changes in the amygdala can occur even later in life and are dependent on timing. Most intriguing is the evidence that effect directions might be reversed dependent on timing, which has important implications for the interpretation of neurostructural alterations. Notably, even if these effects would be due to other differences between the two timing groups (e.g., trauma duration or multiplicity), these opposite effects would still reflect highly relevant nonlinearities as a function of these potential explanatory variables (see limitations for further discussion).

For the more exploratory regions, we did not find any interactions between group and sample, i.e., no indication for the relevance of trauma timing. As would be expected from the meta-analysis which motivated these ROI choices, we found significant main effects of group for all regions except for MTG. Visually, there was a tendency for lower volumes in the PTSD group, especially in the childhood sample. Still, post-hoc tests only confirmed this for the IFOG. Naturally, these non-significant post-hoc tests might be due to the decreased statistical power of the corrected p-values. Hence, while descriptively in line with previous research, we did not find evidence for timing effects in these regions.

We emphasize that the relationship between timing of first trauma experience and brain development is complex and we do not want to create a dichotomy between childhood and adult trauma experience. Many individuals with trauma experience in childhood do also experience aversive events in adulthood. Nevertheless, we think that our study is an important starting point to investigate differences in gray matter volumetry based on first exposure to traumatic experiences beyond childhood and adolescence.

### 4.1 Limitations

The study only included women, which limits the generalizability of results, especially as higher-order interactions between sex, adversity type, and timing have been previously observed (22). Moreover, the studies were cross-sectional and relied on retrospective reports. A longitudinal design could differentiate between brain aberrations as vulnerability factors versus neuroplastic environment-contingent alterations. Still, such studies involve screening of at-risk cohorts with neuroimaging assessments at all time-points, which is thus far only feasible in limited settings, where healthy individuals have a very high prospective probability for trauma exposure, such as military deployment or first-aid workers. Even these studies are dependent on the differential occurrence of changes in psychopathology, a condition not always met (25).

Importantly, we aggregated data from two different studies, one focusing on traumatic experiences during childhood and adolescence and one focusing on adulthood. These studies were conducted at the same facility, using the same scanner, but systematic differences might still occur, for example, due to different recruitment strategies. The samples had notable differences in the severity, type, duration, and multiplicity of traumatic events. Another notable difference is the higher prevalence of comorbid BPD in the childhood sample, which was facilitated by the study procedure. Therefore, it is possible that differences might be attributable to these confounders instead of trauma timing. Still, such differences on confounders might be inherent to realistic occurrences of traumatic events in feasible designs using human neuroimaging. For example, the whole childhood sample experienced maltreatment, a distinct trauma type without a direct counterpart in adulthood which usually coincides with higher multiplicity and duration. Even for singular and highly random adverse events that might seem comparable at first glance (e.g., certain cases of natural disasters and sexual assault), the meaning and impact is vastly different for affected children and adults. Hence, while our design cannot rule out many important confounders, suggesting careful interpretation of results, these confounders might be inherent differences between typical trauma during child- and adulthood. Importantly, the opposite effects for amygdala volume in child- and adulthood are not compatible with a monotonic dose-response effect of variables like duration and multiplicity.

## 4.2 Conclusion

Our findings suggest that amygdala aberrations following adverse experience might be dependent on timing and could occur in response to traumatic events in both child- and adulthood. Adversity effects during child- and adulthood had opposing directions, highlighting the importance to differentiate between neurodevelopmental mechanisms and life-long plasticity. These findings add nuance to the interpretation of brain volumetric associations with adverse experiences. We did not observe such effects of timing for other predefined brain regions implicated in volumetric brain differences related to PTSD. Through our three-group design, our study might inform not only future studies on timing, but also help differentiate effects of psychopathology and trauma-exposure.

## Data Availability

Ethical restrictions to protect participant confidentiality prevent us from making study data publicly available. This also refers to the analysis/experimental code, and any other digital materials, where participant-related information (like sex or psychopathological status) are also included. Readers seeking access to the study data and materials should contact the corresponding author based on a formal collaboration agreement. This formal collaboration agreement indicates that data will be shared with other researchers who agree to work with the authors, and for the sole purpose of verifying the claims in the paper. The data and materials will be released to requestors after approval of this formal collaboration agreement by the local Ethics Committee of the Medical Faculty Mannheim.

https://osf.io/r65uz/?view_only=26f0e011f1d24b279991dff6c57e5fe3

## Disclosures

All authors report no financial relationships with commercial interests.

## Acknowledgement

This study was supported by the German Ministry of Education and Research under Grant No. 01KR1303A and by a grant of the Deutsche Forschungsgemeinschaft (SFB636/C1 to H.F).

## Role of funding sources

All authors declare that the research was conducted in the absence of any commercial or financial relationships that could be construed as a potential conflict of interest.

## Notes

**Author Note**: All authors declare that the research was conducted in the absence of any commercial or financial relationships that could be construed as a potential conflict of interest.

### Competing Interest Statement

The authors have declared no competing interest.

### Author Declarations

The Ethical Review Board of the Medical Faculty Mannheim (Heidelberg University) and the ethics committee of the Goethe University Frankfurt gave ethical approval for this work.

